# Mapping ADHD Heterogeneity and Biotypes through Topological Deviations in Morphometric Similarity Networks

**DOI:** 10.1101/2025.03.27.25324802

**Authors:** Nanfang Pan, Yajing Long, Kun Qin, Isaac Pope, Qiuxing Chen, Ziyu Zhu, Ying Cao, Lei Li, Manpreet K. Singh, Robert K. McNamara, Melissa P. DelBello, Ying Chen, Alex Fornito, Qiyong Gong

**Affiliations:** Huaxi MR Research Center (HMRRC), Department of Radiology, West China Hospital of Sichuan University; Research Unit of Psychoradiology, Chinese Academy of Medical Sciences; Functional & Molecular Imaging Key Laboratory of Sichuan Province, West China Hospital of Sichuan University, Chengdu, China; The Turner Institute for Brain and Mental Health, School of Psychological Sciences and Monash Biomedical Imaging, Monash University, Clayton, Australia; Department of Radiology, Taihe Hospital, Hubei University of Medicine, Shiyan, China; Department of Psychiatry, University of Cincinnati, Cincinnati, USA; Department of Psychiatry and Behavioral Sciences, University of California Davis, Sacramento, USA; Department of Radiology, West China Xiamen Hospital of Sichuan University, Xiamen, China

**Author notes:** **Corresponding Author**: Ying Chen and Qiyong Gong, M.D., Ph.D., Huaxi MR Research Center (HMRRC), Department of Radiology, West China Hospital of Sichuan University, No. 37 Guo Xue Xiang, Chengdu 610041, PR China. Authors contributed equally to this work (Nanfang Pan, Yajing Long).

## Abstract

Attention-deficit/hyperactivity disorder (ADHD) is characterized by considerable clinical heterogeneity. This study investigates whether normative modelling of topological properties derived from brain morphometry similarity networks can provide robust stratification markers for ADHD children. Leveraging multisite neurodevelopmental datasets (discovery: 446 ADHD, 708 controls; validation: 554 ADHD, 123 controls), we constructed morphometric similarity networks and developed normative models for three topological metrics: degree centrality, nodal efficiency, and participation coefficient. Through semi-supervised clustering, we delineated putative biotypes and examined their clinical profiles. We further contextualized brain profiles of these biotypes in terms of their neurochemical and functional correlates using large-scale databases, and assessed model generalizability in an independent cohort. ADHD exhibited atypical hub organization across all three topological metrics, with significant case-control differences primarily localized to a covarying multi-metric component in the orbitofrontal cortex. Three biotypes emerged: one characterized by severe overall symptoms and longitudinally persistent emotional dysregulation, accompanied by pronounced topological alterations in the medial prefrontal cortex and pallidum; a second by predominant hyperactivity/impulsivity accompanied by changes in the anterior cingulate cortex and pallidum; and a third by marked inattention with alterations in the superior frontal gyrus. These neural profiles of each biotype showed distinct neurochemical and functional correlates. Critically, the core findings were replicated in an independent validation cohort. Our comprehensive approach reveals three distinct ADHD biotypes with unique clinical-neural patterns, advancing our understanding of ADHD’s neurobiological heterogeneity and laying the groundwork for personalized treatment.

## 1. Introduction

Attention-deficit/hyperactivity disorder (ADHD) is a common neurodevelopmental disorder characterized by considerable clinical heterogeneity that extends beyond the DSM-5 diagnostic framework[1, 2]. Although its symptoms are typically partitioned into inattentive and hyperactive/impulsive domains, this binary behavioral classification inadequately captures the complexity of ADHD presentations[3]. Clinical observations reveal diverse patterns across DSM-defined domains—with predominantly inattentive individuals showing reduced processing speed, predominantly hyperactive/impulsive individuals exhibiting impaired reactive control[4], and combined presentation demonstrating more severe cognitive deficits[5]—but these superficial distinctions likely oversimplify the diverse neurobiological mechanisms driving ADHD, precluding consensus on more informative and neurobiologically homogeneous subtyping. Supervised approaches (e.g., latent profile analysis) that define subtypes using symptom checklist thresholds typically generate only severity-based cognitive subgroups rather than qualitatively distinct profiles, limiting clinical application that could be derived from prior validation studies[6, 7]. In contrast, data-driven clustering may offer a superior solution by leveraging phenotypic data that best informs how subtypes of disorders should be delineated.

There have been multiple attempts to identify ADHD subtypes by analyzing varying symptom combinations[8], but many of these approaches fail to distinguish normative from atypical variations in phenotypic measures, which may limit their ability to capture clinically meaningful clusters[9]. Normative modeling incorporating neuroimaging metrics offers a strategy for overcoming this limitation[10, 11]. The approach is analogous to pediatric growth charts mapping age-related changes in height and weight, allowing one to quantify centiles of normative variation in a given phenotype that can be used to evaluate the extent to which a given individual deviates from population expectations[12]. Normative modelling thus offers a more powerful framework for understanding atypical features in individuals than classical case-control comparisons, which rely only on contrasts of group means and ignore individual heterogeneity[13].

Previous normative modeling studies in ADHD have primarily examined regional morphology[3], including gray matter volume (GMV)[14–16], cortical thickness[17], and white matter volume[18]. Similarly, large-scale case-control analyses of ADHD also demonstrate brain-wide structural abnormalities[19]. However, regional variations are often coupled across disparate brain systems, and the network-level coupling of deviations from normative regional morphometry remains poorly understood. Morphometric similarity networks (MSN) can be used to characterize the covariance patterns of brain regional features, offering an individualized modeling approach with high robustness and reproducibility for identifying accessible and cost-effective MRI-derived biomarkers[20, 21]. Coupled morphometric variations within MSNs can arise from a combination of inter-regional structural connections through axonal pathways, similarities in cytoarchitecture, and shared patterns of gene expression[21, 22]. Notably, these networks often exhibit hubs—specific regions that are considered particularly central or influential in brain networks by virtue of their coupling patterns[20]. Multiple measures exist for characterizing the “hubness” or centrality of a node within an MSN, indicating that an approach that integrates various information may be particularly fruitful[23]. Such approaches provide a promising framework for understanding brain network alterations in neurodevelopmental disorders[22, 24] and may also serve as neurobiological markers for ADHD[25].

Here, we developed normative models of MSNs in a large multisite cohort of typically developing and ADHD individuals. We specifically focused on measures quantifying the network hubness of a given brain region within the MSN as target phenotypes for normative models, given its importance for neural communication and robustness[26]. Discriminating multivariate components of hubness measures were then used in a semi-supervised clustering analysis to delineate distinct biotypes and their unique clinical and biological profiles, as contextualized with respect to neurochemical (i.e., PET-derived neurotransmitter density maps) and functional (i.e., a large database of task activation maps) correlates. We then cross-validated the putative biotypes identified by this procedure in an independent cohort.

## 2. Results

The overall workflow for our study is depicted in Figure 1.

**Figure 1.**
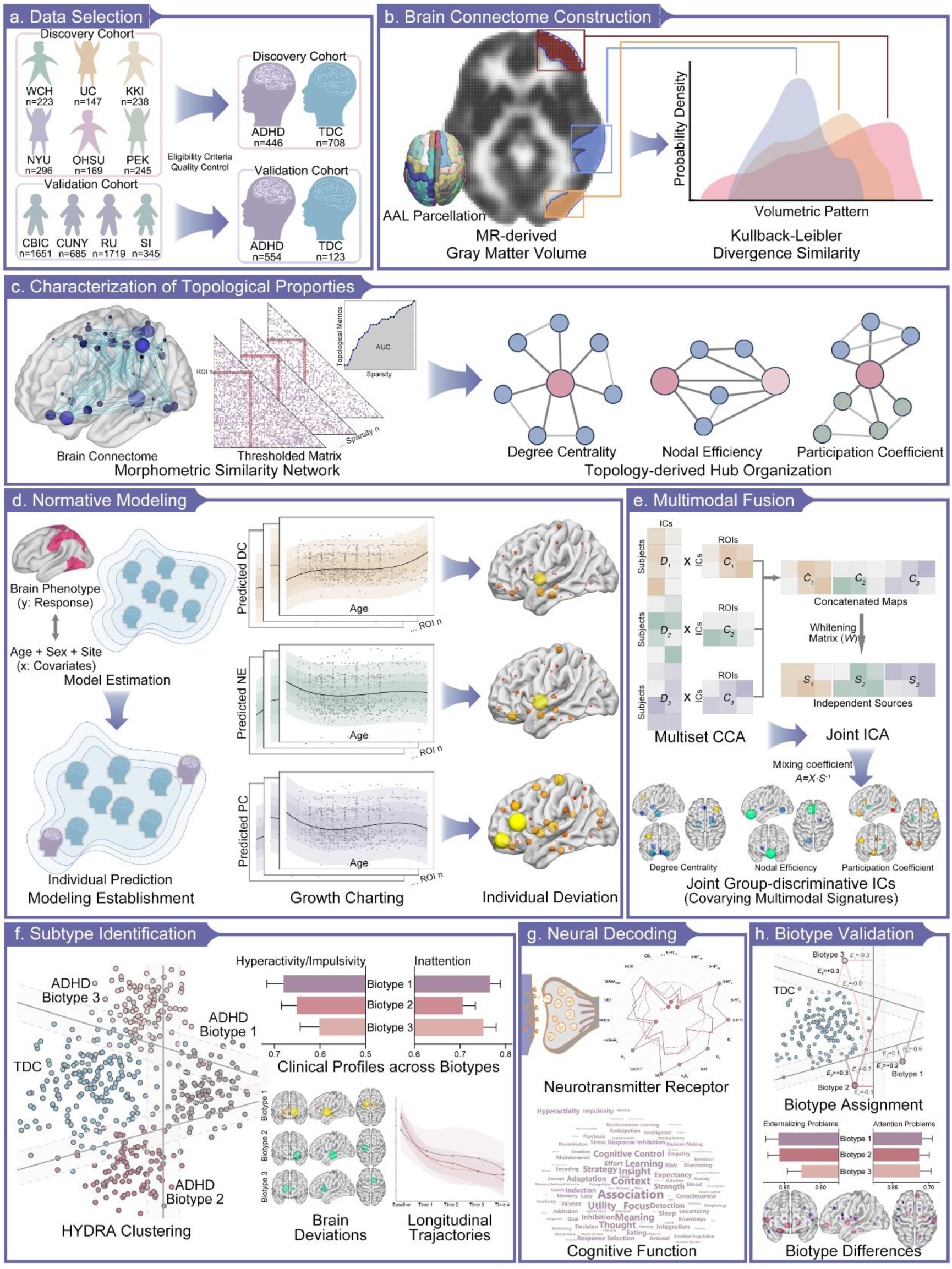
Schematic Overview of Analytical Procedures. (a), Our study comprises two cohorts: a discovery cohort incorporating data from six independent sites (from WCH to PEK, 446 children with ADHD and 708 controls for the final neuroimaging datasets), and a validation cohort derived from the HBN initiative (from CBIC to SI, 554 ADHD cases and 123 controls), both following eligibility assessment and quality control procedures. (b), T1-weighted neuroimages were preprocessed to extract their volumetric patterns of gray matter, which were subsequently parcellated into 90 regions using Automated Anatomical Labeling atlas. Morphological similarity between paired brain regions was quantified using Kullback-Leibler divergence similarity metrics based on probability density function, where higher values indicate stronger morphological similarity between regions and shorter edges in the covariance network. (c), Morphometric similarity networks were constructed across a sparsity range within small-world regime. Network hub organization was comprehensively characterized using three graph-theoretic measures: degree centrality (DC, for local connection density), nodal efficiency (NE, for shortest communication path), and participantion coefficient (PC, for cross-module diversity). (d), Normative models based on Bayesian linear regression were established for each nodal topological phenotype (response variables) to estimate region-specific normative variance, incorporating age, sex, and site effects as covariates. Deviations along normative centiles were characterized using *Z*-scores. (e), mCCA was applied to topological metrics, generating two canonical variates (mixing profile Dk and derived associated map *C_k_*, where *k* is the number of modalities) to capture inter-modal relationships. jICA was then adopted on the concatenated Ck maps to derive maximally independent sources *S_k_* and corresponding whitening matrix W. Individual-wise mixing coefficients (*A_k_*) were calculated to explore the clinical correlations. (f), HYDRA algorithm was implemented to identify potential ADHD biotypes (represented by purple, red, and yellow dots in the simulated model, respectively) based on topological deviations, leveraging linear support vector machines to establish multiple hyperplanes (indicated by gray bands between groups) that simultaneously encompass the control distribution while optimally differentiating ADHD cases. Between-biotype differences in symptom severity, topological deviation patterns, and longitudinal trajectories were exhibited. (g), Biotype-specific topological deviation patterns were examined in relation to neurotransmitter receptor spatial distributions (encompassing 19 distinct receptors). Neurosynth-based meta-analytic task activation maps were used to contextualize our findings, with the relative contributions of cognitive terms visualized through wordcloud plots. (h), Pre-trained HYDRA model was applied to validation cohort to obtain expression scores (*E_i_*, where *i* is the optimal number of biotypes) across dimensions. These scores are computed using the preserved hyperplane weights and bias terms from the training phase, with subsequent cluster assignment based on maximum expression scores.

### 2.1. Clinical Characteristics of Included Data

We included 446 children diagnosed with ADHD (age: mean ± standard deviation: 11.54±2.66, 339 boys, age distributions across sites in Supplementary Figure S1) and 708 typically developing controls (TDC, age: 11.00±2.29, 429 boys) in the discovery cohort and 554 children diagnosed with ADHD (age: 10.09±2.78, 372 boys) and 123 TDC (age: 10.09±2.95, 70 boys) in the validation cohort after eligibility assessment and quality control (selection process detailed in Supplementary Figure S2 and Figure S3). Demographic and clinical characteristics, along with scan parameters, are provided in Supplementary Table S1 and Table S2, respectively. Written informed consent was obtained from all participants and their parents.

### 2.2. Individual Deviations on Topological Properties

Anatomic T1-weighted images of included samples were analyzed to generate individual GMV maps and the Automated Anatomical Labeling atlas was adopted to segment each brain’s gray matter into 90 regions[27] (see Methods, Section 4.2). MSNs were constructed using Kullback-Leibler divergence similarity (KLS) metric. Since individual topological metrics may bias hub estimations according to unique assumptions about network information flow, and no single method definitively detects hubs, a multi-metric approach is essential to robustly identify hubs that exhibit diverse connectivity properties[23]. Three topological metrics of DC, NE, and PC were calculated to assess each region’s MSN hubness in terms of local connection density, shortest communication path, and cross-module diversity, respectively[28, 29] (see Methods, Section 4.4).

For each of the three hubness metrics and each brain region, we used a normative model with Bayesian linear regression to estimate centiles of normative variation in a training set comprising only typically developing individuals. Individual deviations from these normative expectations were then quantified via *Z*-scores. Our normative models demonstrated satisfactory fitting performance for all three topological metrics as illustrated in Supplementary Figure S4, with site effects presented in Supplementary Table S3. To summarize the nodal heterogeneity of extreme deviations in each group, the deviations of topological patterns were thresholded at |*Z*|≥2.0 for visualization and interpretation[30, 31]. Among children with ADHD, 95.07%, 96.41%, and 71.75% of them exhibited at least one extreme deviation in DC, NE, and PC from the normative range, respectively (distributions of extreme positive and negative deviations are presented in Supplementary Figure S5). We then computed the proportion of children with suprathreshold positive or negative deviations (i.e., atypically increased or reduced phenotypes with |Z|≥2.0) at each topological node, revealing distinct patterns across topological metrics (Figure 2a).

**Figure 2.**
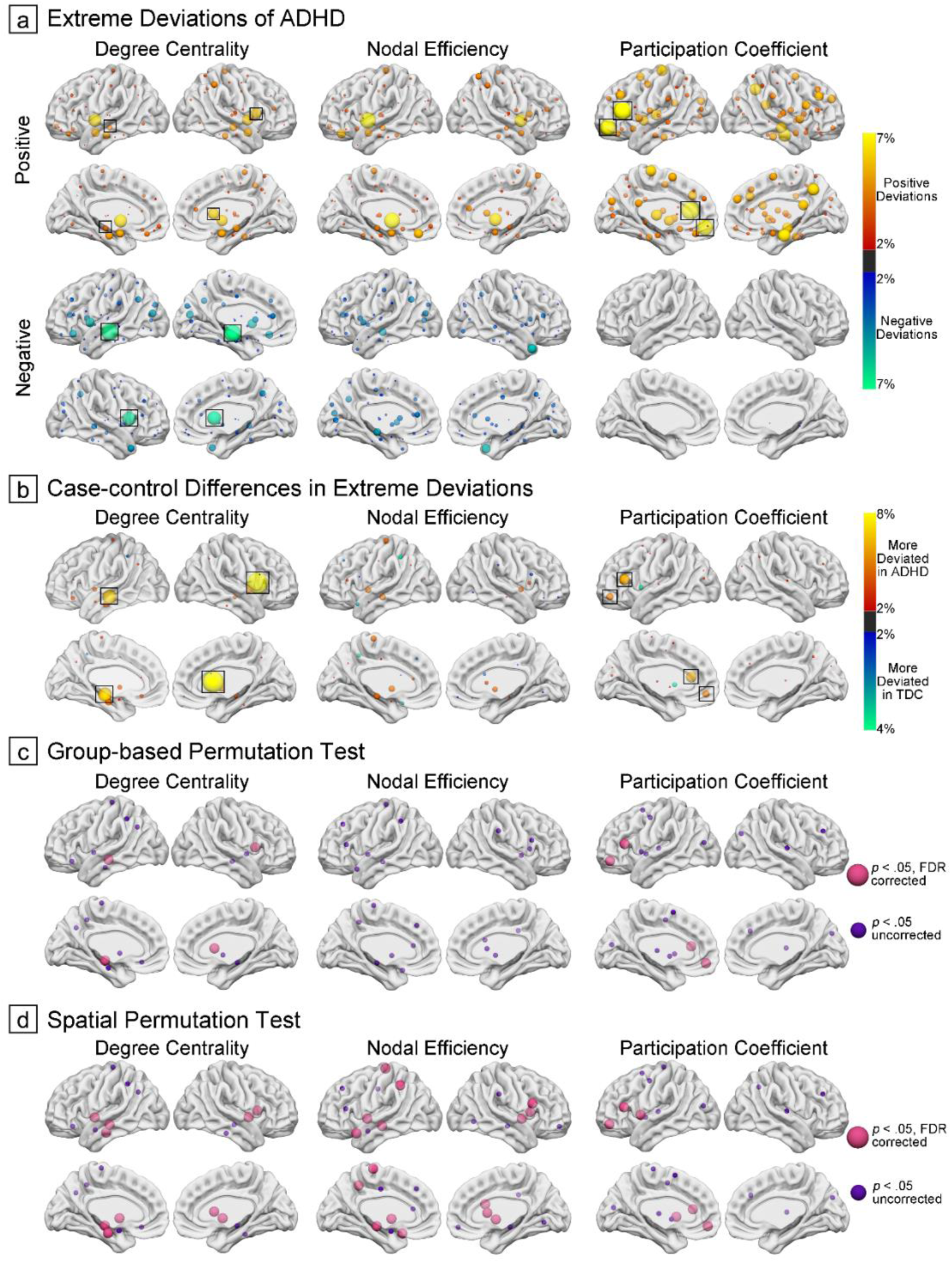
Nodal Heterogeneity of Extreme Topological Deviations. (a), Proportion of subjects displaying extreme topological deviations across brain nodes. Positive deviations are depicted in red-yellow, and negative deviations in blue-green, shown separately for each topological metric. Nodes that survived both group-based and spatial permutation tests were highlighted with black square. (b), Differential patterns between groups, computed by subtracting TDC overlap maps from ADHD overlap maps. Red-yellow indicates nodes with higher deviation prevalence in ADHD, while blue-green represents nodes with higher deviation prevalence in TDC. (c) and (d), Nodes exhibiting significant case-control differences in extreme deviation patterns, assessed by both group-based and spatial permutation tests. Nodes exceeding statistical thresholds are highlighted.

Case-control comparison at the level of extreme deviation overlap within each region (Figure 2b-d and Supplementary Table S4) revealed pronounced DC differences in the caudate and hippocampus (observed difference=7.79% and 6.33%, respectively; false discovery rate (FDR)-corrected *p*<.001 in both group-based and spatial permutation testing), and PC differences were primarily scattered in the inferior frontal gyrus and orbitofrontal cortex (observed difference=5.50% and 4.52%, respectively; *p*_FDR_≤.045 in all permutation testing). For NE, significant spatial overlap patterns emerged in the hippocampus and pallidum (observed difference=4.31% and 4.24%, respectively), showing significance in spatial permutation testing (both *p*_FDR_<.001) but more modest effects in group-based analysis (*p*_FDR_=.114 and .180, respectively).

### 2.3. Covarying Multimodal Signatures

To identify joint independent components that encompassed co-varying multi-metric patterns, topological phenotypes were subjected to a data-driven model—multiset canonical correlation analysis plus joint independent component analysis (mCCA + jICA). During this analysis, topological metrics were projected onto two canonical variates to delineate spatial maps and generate independent sources (see Methods, Section 4.7)[32]. The optimal selection of eight components preserved a substantial proportion of the explained variance across metrics (80.7%, 83.2%, and 82.9% for DC, NE, and PC, respectively). Notably, the fusion model yielded a joint component that demonstrated significant case-control differences in mixing coefficients across topological metrics (Figure 3a, *t*=-2.49, *p*=.013; *t*=-2.24, *p*=.025; and *t*=2.19, *p*=.029, respectively). Within this group-discriminative component, covarying patterns were predominantly localized to the rectus gyrus, within the orbitofrontal cortex, (Figure 3b and Supplementary Table S5; significant observations in all three topological modalities, *Z*=-2.11, -3.15, and 2.31, respectively). Further analysis revealed significant associations between hyperactivity/impulsivity symptoms and mixing coefficients in three modalities in ADHD children (Figure 3c, *r*=0.12, *p*=.013; *r*=0.11, *p*=.021; and *r*=-0.10, *p*=.040, respectively). However, no significant linear relationships were detected between these coefficients and inattention scores.

**Figure 3.**
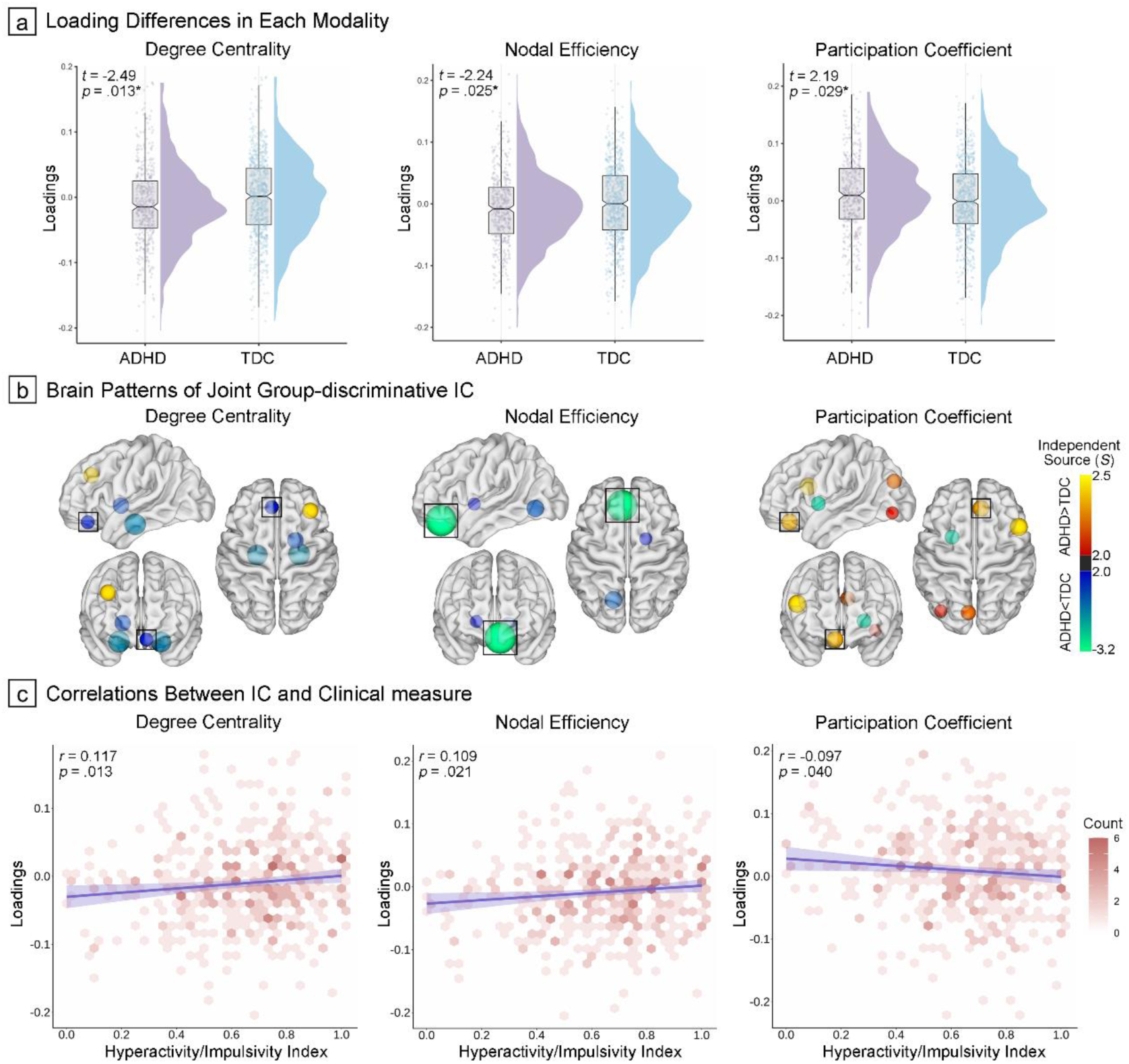
Covarying Topological Signatures Distinguishing ADHD from TDC (a) Raincloud plots illustrating the distribution of component loadings across modalities and groups, with statistical comparisons indicated in the upper left corner. (b) Spatial distribution of the independent component (IC) based on independent source (*S*). Red-yellow nodes indicate ADHD>TDC, while blue-green nodes indicate the opposite pattern. Consistent alterations across all three topological properties were highlighted with a black square. (c) Density scatter plots depicting associations between component loading values and clinical profiles.

### 2.4. ADHD Biotypes Corresponding to Clinical Profiles

Heterogeneity through discriminative analysis (HYDRA), a semi-supervised algorithm, was implemented to analyze individual deviations (*Z*-scores from normative ranges) in three hubness measures to identify putative biotypes in ADHD children[33, 34]. The algorithm uses support vector machines to create multiple hyperplanes that separate controls from cases to form a convex polytope, with each facet representing a distinct biotype (See Methods, Section 4.7). The three-biotype solution best captured the heterogeneity of topological deviations in our models and this scheme showed high robustness and reliability according to four criteria. First, this clustering solution yielded the highest adjusted Rand index (ARI, 0.21 for *k*=3, 0.18 for *k*=2 and 0.17 for *k*=4). Second, in permutation testing, ARIs at *k*=3 were significantly higher than null expectations (ARI_alternative_ vs. ARI_null_ = 0.19 vs. 0.17, *t*=3.42, *p*<.001, Supplementary Figure S6). Third, when excluding females, the neural deviation patterns were similar compared to the full-sample model, supporting its robustness (Supplementary Figure S7). Finally, *k*=3 also emerged as the optimal solution in split-half cross-validation based on ARIs (Split 1: 0.21±0.04, Split 2: 0.20±0.04, ARIs for other cluster solutions provided in Supplementary Figure S8), with the clustering scheme demonstrating replicability in neural patterns (Supplementary Figure S9).

Biotype 1, 2, and 3 consisted of 142, 177, and 127 ADHD children, respectively (see their distribution patterns in DSM profiles in Supplementary Figure S10). Kruskal-Wallis analysis of clinical manifestations revealed distinct patterns across the three ADHD biotypes, with significant differences across putative biotypes in both inattention (Figure 4a, *H*=8.94, *p*=.011, measured by Connors’ Rating Scale or ADHD Rating Scale) and hyperactivity/impulsivity (*H*=8.35, *p*=.015). Specifically, Biotype 1 exhibited the most severe symptomatology, with the highest scores in both inattention (0.77±0.14) and hyperactivity/impulsivity (0.68±0.22). In contrast, Biotype 2 displayed predominantly hyperactivity/impulsivity (0.65±0.23) accompanied by moderate inattention (0.71±0.19). Biotype 3 manifested a distinctive profile characterized by pronounced inattention (0.75±0.16) coupled with relatively milder hyperactivity/impulsivity (0.60±0.24). Post-hoc comparisons identified significant differences in hyperactivity/impulsivity between Biotypes 1 and 3 (*t*=2.87, *p*_adjusted_=.012) and inattention between Biotypes 1 and 2 (*t*=2.81, *p*_adjusted_=.015).

**Figure 4.**
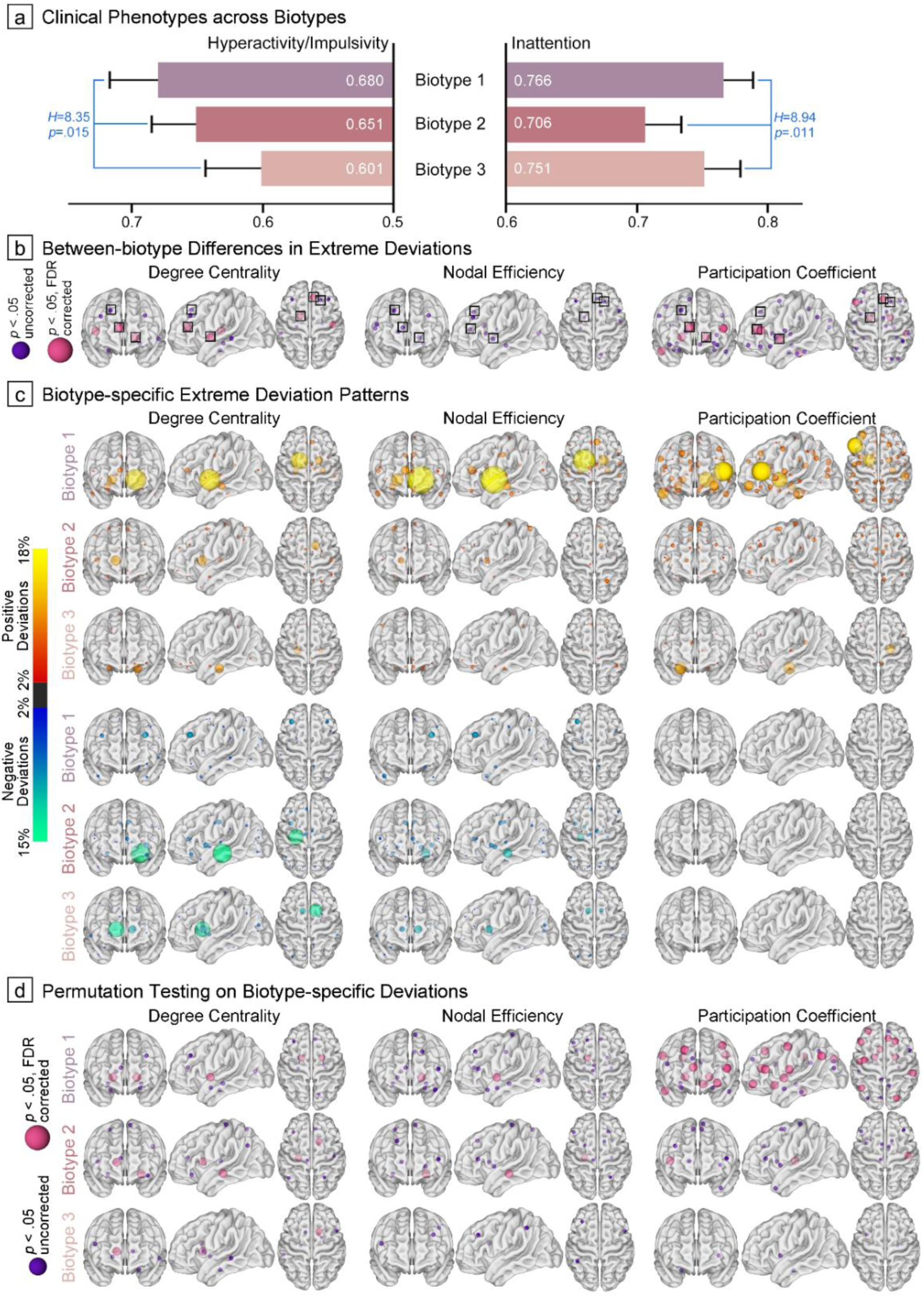
ADHD Biotype Identification Based on HYDRA Modeling. (a) and (b), Between-biotype differences in symptom severity and atypical neural mechanisms. Neuroanatomical locations of hub regions are shown across topological modalities, with smaller purple nodes indicating suprathreshold significance at *p*_uncorreced_<.05 and larger pink nodes indicating suprathreshold significance at *p*_FDR_<.05. Consistent alterations across all three topological properties were highlighted with a black square. (c), Biotype-specific extreme deviation maps were generated by computing the proportion of subjects with suprathreshold deviations. Red-yellow nodes represent atypically increased topological metrics, while blue-green nodes indicate decreased metrics. Nodes exhibiting significant between-biotype differences across all modalities were highlighted with black squares, in accordance with our proposed segregation model. (d), Hub region nodes that demonstrated statistical significance in group-based permutation tests when compared to null patterns.

When examining nodal between-biotype differences (Figure 4b), the right anterior cingulate cortex (χ²≥12.16, *p*≤.002), left pallidum (χ²≥11.33, *p*≤.003), and right superior frontal gyrus (χ²≥7.75, *p*≤.021) demonstrated consistent alterations across all three topological metrics. However, *p*-values for NE did not survive FDR correction (see Supplementary Table S6 for details and modality-specific differences). Relative to other biotypes, Biotype 1 exhibited the most extreme deviations across the above nodes and all metrics, particularly in PC (6.34%, 13.38%, and 9.15%, respectively). Biotype 2 instead demonstrated more atypical patterns of DC in the left pallidum (7.34%) and NE in the right anterior cingulate cortex (9.04%), while Biotype 3 showed greater alterations in the right superior frontal gyrus (7.09% and 8.70% in DC and NE, respectively). In comparison with the normative range derived from TDCs (Figure 4c-d, details in Supplementary Table S7), Biotype 1 showed consistently significant positive deviations in the left pallidum across three topological metrics (extreme positive deviations≥13.38%, *p*_FDR_<.001). Biotype 2 showed positive deviations in the right caudate (extreme positive deviations≥6.78% and *p*≤.014 across modalities, with only differences in DC surviving FDR correction), whereas Biotype 3 exhibited widespread deviations in frontostriatal substrates, though these alterations were inconsistent across the topological metrics.

Analysis of 4-year longitudinal trajectories in those medication-naïve ADHD based on the Child Behavior Checklist revealed no significant time-by-biotype interaction effects (Supplementary Table S8, all |*Z*|<2.0, *p*>.05) for both attention and externalizing problems, suggesting parallel symptom improvement (time effects: all |*Z*|≥5.41, *p*<.001) in these two domains across biotypes. Intriguingly, when examining deficient emotional self-regulation—a key source of heterogeneity linked to elevated psychopathology risk in late-adolescent ADHD[35]—Biotype 1 showed more persistent symptoms compared to the marked decreases observed in Biotypes 2 and 3 (longitudinal trajectories depicted in Supplementary Figure S11; time-by-biotype interactions: Biotype 1 vs. 2: *Z*=-2.11, *p*=.035; Biotype 1 vs. 3: *Z*=-2.66, *p*=.008). We also tracked the development of mood disorders during follow-up: while Biotype 1 showed a higher rate of mood disorder comorbidity (25.0%) compared to Biotypes 2 and 3 (9.8% and 5.0%, respectively), these differences were not statistically significant (*χ²*=2.89, *p*=.219).

### 2.5. Biotype Decoding

We investigated the molecular bases of our topology-derived ADHD biotypes by examining their associations with neurotransmitter receptor distributions[36]. Drawing from a comprehensive dataset encompassing 1,308 healthy participants in 31 different PET studies, we acquired 19 receptor density maps categorized into nine neurotransmitter systems, as collated in the neuromaps database[37]. Our investigation of spatial relationships between these neurotransmitter density maps and fused topological deviations across biotypes (See Methods, Section 4.9, Supplementary Figure S12) revealed that fused topological hub abnormalities in Biotype 1 of ADHD exhibited significant spatial correspondence with the serotonin (Figure 5a, 5-HT_4_, *r*=0.37, *p*_spin-FDR_=.003; 5-HTT, *r*=0.37, *p*_spin-FDR_=.005), dopamine (D_2_, *r*=0.25, *p*_spin-FDR_=.032), acetylcholine (α_4_β_2_, *r*=0.26, *p*_spin-FDR_=.032; M_1_, *r*=0.41, *p*_spin-FDR_<.001), and histamine (H_3_, *r*=0.29, *p*_spin-FDR_=.020) neurotransmitter density distributions. The atypical patterns of Biotype 2 showed significant anticorrelations with glutamate (mGluR_5_, *r*=-0.24, *p*_spin-FDR_=.030), cannabinoid (mGluR_5_, *r*=-0.37, *p*_spin-FDR_<.001), and serotonin (5-HT_1A_, *r*=-0.30, *p*_spin-FDR_<.001; 5-HT_2A_, *r*=-0.34, *p*_spin-FDR_<.001) systems, while Biotype 3 only exhibited significant anticorrelations with one serotonin receptor (5-HT_2A_, *r*=-0.25, *p*_spin-FDR_=.032). Spatial correlations with individual topological modalities were similar with the fusion model as presented in Supplementary Figure S13.

**Figure 5.**
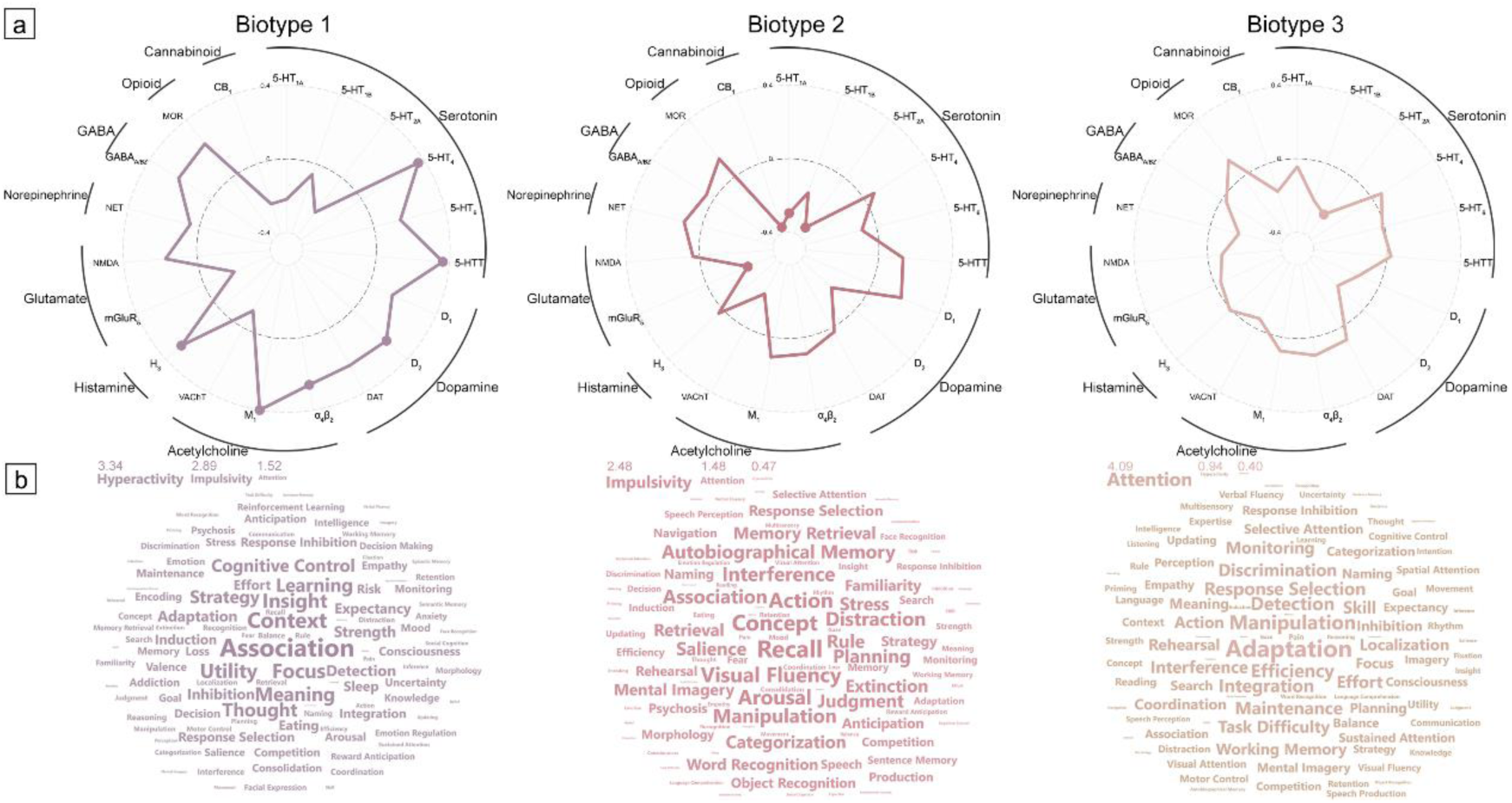
Neurotransmitter and Cognitive Profiles of Biotype-specific Topological Deviations. (a), Spider plot illustrating the spatial correlations between maps of integrating topological deviations and neurotransmitter receptor density distributions across biotypes. Each axis represents a distinct receptor subtype, with outer circles indicating broader neurotransmitter systems. Distance from the dashed baseline reflects correlation strength. Biotypes are distinguished by color coding. Solid dots indicate statistically significant correlations after correction for spatial autocorrelation using spin tests (*p*_spin_<.05). (b), Wordcloud visualization of cognitive terms derived from Neurosynth meta-analyses, scaled by their respective Z-scores of PLS1 loading across biotypes. Word size is proportional to absolute Z-score magnitude. Core ADHD clinical dimensions (attention, hyperactivity, and impulsivity) are highlighted at top of the wordcloud.

To validate the ADHD biotypes’ clinical profiles and broaden our understanding beyond the behavioral checklists, we additionally examined associations between each biotype and psychological processes using Neurosynth-derived meta-analytic task-based activation maps[38]. Partial least squares (PLS) analysis between biotype-specific fused topological deviations and Neurosynth meta-analytic maps identified components that capture the highest explained variance across maps of cognitive terms, accounting for 12.7%, 9.6%, and 8.5% of variance for Biotypes 1-3, respectively (*p*_spin_=.010, .043, and .048, and see Supplementary Figure S14 for their spatial correlations with fused topological deviations). The contribution of cognitive terms to PLS components across biotypes was hierarchically organized based on absolute *Z* scores of PLS loadings in bootstrapping (Figure 5b and Supplementary Table S9). Notably, Biotype 2 predominantly exhibited the “impulsivity” feature (|*Z|*=2.48), with relatively modest contributions from “attention” (|*Z|*=1.48) and “hyperactivity” (|*Z|*=0.47) domains. In contrast, Biotype 3 demonstrated a pronounced “attention” pattern (|*Z|*=4.09), while showing minimal association with “hyperactivity” (|*Z|*=0.94) and “impulsivity” (|*Z|*=0.40) terms. Biotype 1 displayed a more balanced expression of all three terms, with comparable magnitudes across “hyperactivity” (|*Z|*=3.34), “impulsivity” (|*Z|*=2.89), and “attention” (|*Z|*=1.52) terms. Together, these findings indicate a strong correspondence between biotype-specific clinical features and their associated cognitive patterns.

### 2.6. Biotype Validation

In validating our model’s generalizability, the pre-trained HYDRA clustering solution identified three ADHD biotypes in the validation cohort, with 173, 204, and 177 children assigned to Biotypes 1, 2, and 3, respectively. Significant differences in the hyperactive/impulsive domain between biotypes (assessed by Conners’ Rating Scale, *H*=6.58, *p*=.037) maintained consistency with the discovery cohort, showing a decreasing severity profile from Biotype 1 to 3 (Figure 6a; 0.71±0.14, 0.69±0.14, and 0.67±0.14, respectively). Post-hoc comparisons demonstrated significant differences between Biotypes 1 and 3 (*t*=2.55*, p*_adjusted_*=*.032). However, the inattention profiles showed no significant between-biotype differences (*H*=4.14, *p*=.126). Due to the substantial missing data in the Conners rating scale (25.3%), we also utilized the Child Behavior Checklist (0.4% missing data) to validate our clustering findings. Complementary analyses also corroborated distinct patterns of hyperactivity/impulsivity (*H*=7.19*, p=*.027; 0.658±0.12, 0.657±0.12, and 0.626±0.13 for Biotypes 1 to 3; between Biotypes 1 and 3: *t*=2.48, *p*_adjusted_=.040). While patterns of attention problems in the validation cohort paralleled those of the discovery sample (0.691±0.09, 0.687±0.10, and 0.688±0.10 for Biotypes 1 to 3), the differences did not reach statistical significance (*H*=0.49, *p*=.783).

**Figure 6.**
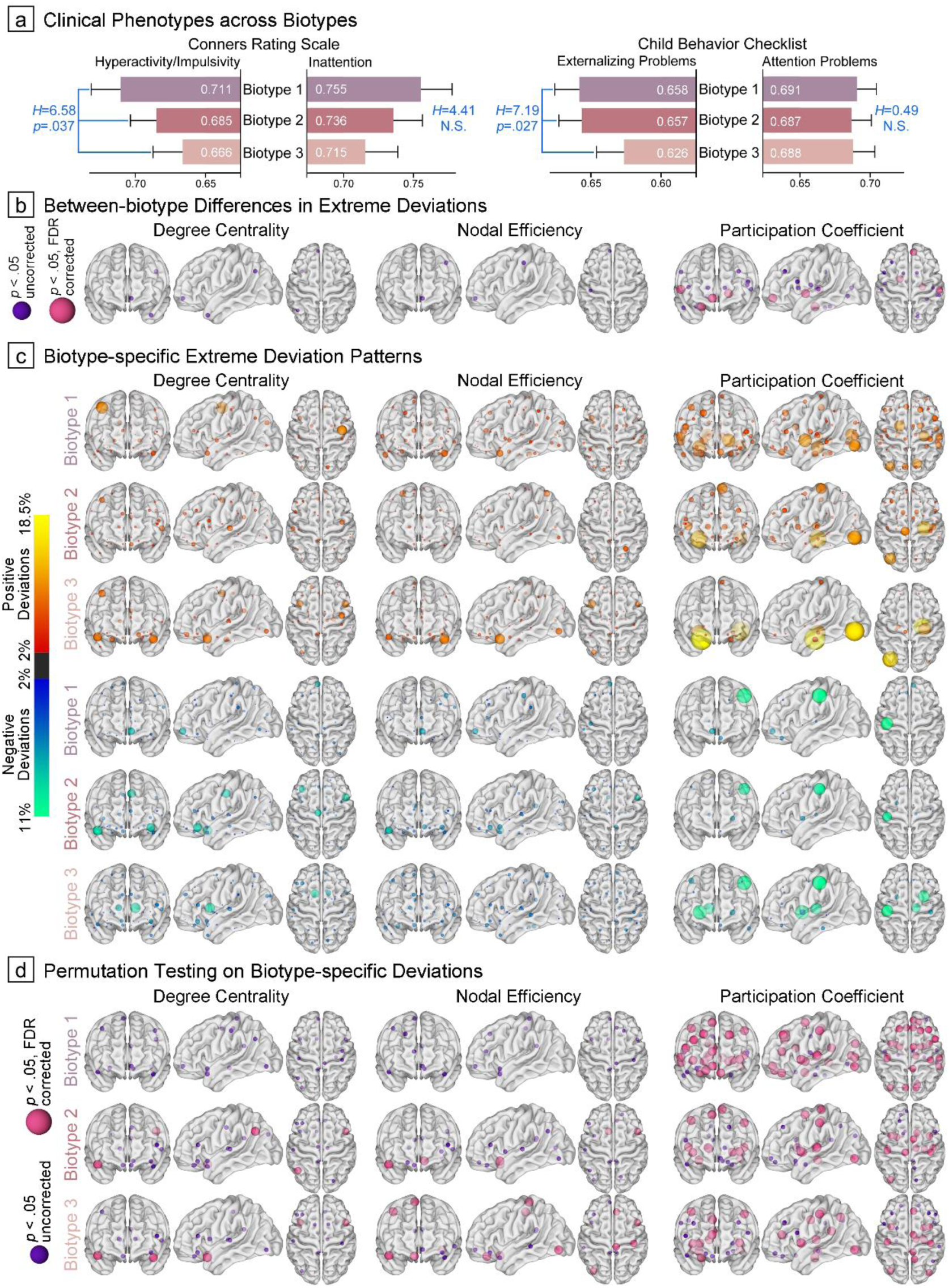
Biotype Validation Based on Pre-trained HYDRA Model. (a) and (b), Between-biotype differences in symptom severity and atypical neural mechanisms. Neuroanatomical locations of hub regions were exhibited across topological modalities, with smaller purple nodes indicating significance at *p*_uncorreced_<.05 and larger pink nodes indicating significance at *p*_FDR_<.05. (c), Biotype-specific extreme deviation maps were generated by computing the proportion of subjects with suprathreshold deviations. Red-yellow nodes represent atypically increased topological metrics, while blue-green nodes indicate decreased metrics. (d), Hub region nodes that demonstrated statistical significance (*p*_uncorreced_<.05 or *p*_FDR_<.05) in group-based permutation tests when compared to null patterns.

Biotype-specific topological deviation patterns showed significant consistency between training and validation cohorts, as evidenced by high correlations between their averaged feature vectors (*r*_Biotype1_=0.72, *r*_Biotype2_=0.80, *r*_Biotype3_=0.76; all *p*<.001). The most pronounced between-biotype differences emerged with respect to PC deviations (16 significant areas), paralleling observations from the discovery cohort. The hub organization of orbitofrontal cortex varied between biotypes across all three metrics (χ²≥8.11, *p*≤.017, proportions of extreme deviations progressively decreasing from Biotype 1 to 3 (Figure 6b and Supplementary Table S10). Regarding biotype-specific neural bases (Figure 6c-d), Biotype 1 demonstrated atypically positive hub organization in structural networks across topological metrics, with particularly pronounced PC deviations compared to controls (detailed in Supplementary Table S11), while Biotypes 2 and 3 displayed more distributed negative deviations in both DC and PC measures.

## 3. Discussion

Phenotypic heterogeneity poses a major challenge in the diagnosis and treatment of ADHD[7], as the current diagnostic framework assigns a single diagnostic label to what is fundamentally a heterogeneous syndrome that likely arises from diverse neural mechanisms[39]. Using topological normative modeling, we identified neural deviation patterns in ADHD children on an individual basis, yielding a covarying component derived from multimodal patterns. Subsequent HYDRA modeling uncovered three distinct biotypes with unique clinical and biological profiles, longitudinal symptom trajectories, and molecular signatures. Both cognitive patterns from the large-scale Neurosynth database and validation in an independent validation cohort largely replicated the clustering findings observed in the discovery datasets. Our study advances the understanding of ADHD heterogeneity through a novel hub-oriented fusion framework that integrates multimodal topological deviations in morphometric similarity networks. The replication of our findings across two cohorts suggest that our approach may offer a feasible framework for stratifying ADHD cases.

### 3.1. Topological Deviations of ADHD from Normative Centiles

Compared to the normative centiles of TDCs, ADHD was associated with distinct patterns of extreme deviation that depended on the topological metric: DC and NE exhibited pronounced differences in subcortical substrates, particularly the caudate, pallidum, and hippocampus, whereas PC abnormalities emerged in the ventral prefrontal cortex, specifically its inferior and orbital sections. Network hub dysfunction can significantly impact network efficiency due to altered gray matter composition and metabolism[26, 40]. Our finding that children diagnosed with ADHD exhibit prominent DC and NE deviations in subcortical hubs—particularly the striatum—suggests a weakened functionality consistent with ENIGM-ADHD consortium findings of reduced subcortical GMV[41]. Through extensive striatal-hippocampal connections[42], hippocampal hub function is similarly compromised underlying abnormal maturation of the limbic system[43]. Given the altered PC patterns, while the inferior frontal gyrus and orbitofrontal cortex typically serve as provincial hubs mediating within-module communication in normative ranges[44], they may shift to connector hubs facilitating global inter-modular integration in ADHD. Increased global information processing in these regions may disrupt neural information integration, leading to dedifferentiation—the breakdown of specialized neural processes[26]. Alternatively, this shift may represent compensatory adaptation to reduced processing capacity in other nodes, manifesting as early increases in hub functionality of the inferior frontal gyrus and orbitofrontal cortex.

The classical ADHD model of pathophysiology suggests delayed brain maturation[19, 45], particularly in frontal cortex and frontostriatal connections[43]. Within the frontostriatal circuit underlying reward and motivation, ventral prefrontal regions regulate striatal activity with striatal feedback modulating prefrontal control signals[46], while the pallium transfers information to the thalamus[47]. Striatal inhibitory projections modulate dopaminergic neuron activity through membrane current regulation[42], which is fundamental to synaptic plasticity. Altered frontostriatal connectivity may thus explain our observed striatal alterations in MSN hub organization[48]. Prefrontal areas, particularly the inferior frontal gyrus[49], and their associated networks could act as a brake for inhibitory control and decision-making optimization, while striatal substrates drive impulsivity[50, 51]. Frontostriatal disruptions in ADHD compromise executive control, reward processing, and value-based decision-making, manifesting as executive function deficits. Therapeutically, psychostimulants may normalize striatal neural responsiveness and connectivity metrics in children with ADHD[52, 53].

Applying multi-metric approach to ADHD may reveal previously undetectable patterns of deviation from normative hub organization[54], potentially enabling robust quantification and prediction of cognitive performance through subsequent data mining and model generalization[32]. Our multimodal fusion analysis then yielded a joint component that recapitulates ADHD-related atypical abnormalities, with covarying patterns predominantly localized within the orbitofrontal cortex that correlated with hyperactivity/impulsivity symptoms. This finding aligns with the orbitofrontal cortex’s role of orchestrating the transition between impulsive and reflective behaviors while integrating reward signals with goal-directed actions[55, 56]. Aligned with our findings, neuroimaging studies of ADHD have extensively revealed alterations in the orbitofrontal cortex, which plays a crucial role in reward and motivation processing through its interactions with striatal circuits[46, 47].

### 3.2. Characterizations of Distinct Biotypes

Our data-driven clustering approach to identifying neurobiological ADHD subtypes offers advantages over traditional clinical classifications. This method parses ADHD’s biological heterogeneity beyond DSM-based subtypes—their underlying biological mechanisms may be more complex than these clinical classifications suggest[6], potentially uncovering novel and real-existing subtypes that clinical assessments may miss. Moreover, it mitigates referral bias-induced sample size imbalances while providing a foundation for developing personalized treatment strategies specific to each biotype[57, 58]. This neurobiologically-informed multi-metric classification shows promise in identifying more homogeneous and clinically relevant ADHD subtypes[3].

The three biotypes that emerged from HYDRA modeling are characterized by severe overall symptoms (Biotype 1), predominant hyperactivity/impulsivity (Biotype 2), and marked inattention (Biotype 3). Biotype 1 showed stronger MSN hub deviations in three metrics across anterior cingulate cortex, superior frontal gyrus, and pallidum, while Biotypes 2 and 3 displayed distinct alterations in DC and NE—Biotype 2 exhibited atypical patterns in the anterior cingulate cortex and pallidum, and Biotype 3 in the superior frontal gyrus. Consistently, a task-based fMRI study identified parallel and distinct neural dysfunctions in ADHD, linking response inhibition deficits to anterior cingulate-insular alterations and attention deficits to superior frontal gyrus changes[59]. In the action-mode network[60], the anterior cingulate cortex orchestrates flexible cognitive control transitions and action monitoring through circuits with the insula[61], while the pallidum modulates frontostriatal inhibitory inputs[62]. These disruptions may disrupt the fine-tuned balance of inhibitory control, contributing to executive dysfunction in ADHD[19, 41]. Conversely, the superior frontal gyrus, a key component of the default mode network, shows dysregulation in ADHD that may impair sustained attention through interference hypothesis[3].

Beyond core ADHD symptoms, these biotypes demonstrated distinct longitudinal trajectories in deficient emotional self-regulation, representing a crucial source of clinical heterogeneity[63]. Notably, Biotype 1 exhibited more persistent symptoms and a higher rate of mood disorder conversion compared to the marked symptomatic improvement observed in other biotypes. The impact of emotional dysregulation on ADHD development also manifests through disrupted executive functioning, spanning from preschool through early school years[63]. Understanding these cross-domain interactions is essential for integrating cognitive and emotional frameworks of ADHD and identifying psychopathologically vulnerable individuals[64]. Further, these biotype-specific biomarkers may hold prognostic value, particularly for Biotype 1 which shows a more severe developmental trajectory requiring prompt and sustained intervention[65].

### 3.3. Contextualization beyond Biotypes

The MSN hub profiles of the biotypes identified here exhibited distinct spatial patterns of correspondence with neurotransmitter receptor distributions: Biotype 1 exhibited widespread abnormalities across dopaminergic, serotonergic, cholinergic, and histaminergic systems, Biotype 2 showed selective alterations in glutamate, cannabinoid, and serotonergic systems, while Biotype 3 displayed confined abnormalities primarily in the serotonergic system. For neurochemical profile of Biotype 1, dopaminergic neurons, which display tonic spontaneous firing patterns regulated by inhibitory striatal projections[42], interact with the serotonergic system through orbitofronto-striatal circuits to modulate hyperactivity/impulsivity in ADHD[66]. Their interplay is evidenced by the efficacy of both dopamine reuptake inhibitors like methylphenidate and selective serotonin reuptake inhibitors[66, 67]. Cholinergic dysfunction within the prefrontal cortex manifested prominently in the combined ADHD subtype—an observation consistent with our findings[68, 69], particularly affecting motor control and attention. Emerging evidence also implicated neuroinflammation as a key mechanism in ADHD pathophysiology[70], notably through altered histaminergic distribution patterns. Understanding these complex neurochemical interactions mentioned above presents promising opportunities for developing targeted treatments for the most severe ADHD biotype. In hyperactivity/impulsivity-dominant Biotype 2, alterations in glutamate systems may reflect disrupted excitatory neurotransmission within caudate-related circuits, which mediate cognitive and affective processes including reward processing, supported by both pharmacological and genomic studies[71]. A randomized controlled trial has demonstrated significant improvement in hyperactivity/impulsivity symptoms with cannabinoid treatment[72]. As ADHD individuals transition toward adulthood, inattention symptoms tend to persist and are often accompanied by increased vulnerability to emotional dysregulation, suggesting that prescriptions for Biotype 3 targeting the serotonergic system may provide additional clinical benefits[73].

Our Neurosynth-decoding findings demonstrated robust alignment between biotype-dominant clinical profiles (i.e., inattention and hyperactivity/impulsivity) and expression patterns of their corresponding cognitive terms. This correspondence to a large-scale database validates our ADHD biotype classification system derived from topological deviations. Further, the application of a pre-trained model in the validation cohort substantially replicated the clustering patterns observed in the discovery datasets, offering external validation for our proposed subtyping. Our integrated approach— encompassing explicit conceptual models, assumptions, and goals—adheres to the recommended workflows for investigating ADHD heterogeneity, progressing systematically from feature selection and reduction through data-driven clustering to external validation[6, 65]. This comprehensive and rigorous approach therefore highlights a potential for advancing both mechanistic understanding and targeted treatments for ADHD.

### 3.4. Limitation

While our findings offer a new characterization and subtyping of ADHD’s heterogeneity, we do not claim that they are exhaustive due to following limitations. First, our samples were not medication- or comorbidity-naïve. Although previous mega-analysis research suggests that psychostimulant use and comorbid conditions do not significantly influence brain morphometric findings in ADHD[41, 74], we cannot completely rule out their potential confounding effects. Second, the overlap in topological deviation patterns between ADHD and TDCs underscores the subtle and complex nature of ADHD-related brain alterations. This complexity may explain why pathognomonic biomarkers remain elusive[15], reflecting the inherent challenges in identifying discrete neurobiological markers for this disorder. Third, it is unclear whether the observed biotype differences can be fully explained by severity across dimensions, given that our data-driven approach was designed to identify novel patterns not captured by traditional clinical measures[6]. Future research should focus on assessing whether these biotype classification models have clinical utility, particularly through the development of decision-making algorithms with optimal cutoff scores for targeted treatments.

### 3.5. Conclusion

We have identified three distinct topology-derived biotypes, each characterized by unique clinical-neural profiles, longitudinal trajectories, and molecular signatures. The robustness of these biotypes, validated through cognitive profiles from both large-scale databases and external cohorts, provides compelling evidence for an objective and reliable basis for parsing ADHD heterogeneity in a clinically meaningful way. Our comprehensive approach, from feature extraction through data-driven clustering to external validation, offers a promising framework for parsing the inherent clinical heterogeneity of an ADHD’s diagnosis, which may ultimately create a path toward developing personalized therapeutic strategies.

## 4. Methods

### 4.1. Data Collection

Our discovery project analyzed multisite data from six independent datasets (Figure 1a): West China Hospital of Sichuan University (WCH), University of Cincinnati (UC), Kennedy Krieger Institute (KKI), New York University Langone Medical Center (NYU), Oregon Health & Science University (OHSU), and Peking University Institute of Mental Health (PKU). Among these, KKI, NYU, OHSU, and PKU datasets were retained and derived from both the ADHD-200 and ABIDE initiatives after excluding sites that had fewer than ten subjects per group or did not use 3.0 Tesla scanners (inclusion and exclusion criteria across sites provided in supplementary Table S12)[75]. To reconcile sample and cross-site heterogeneity, we excluded left-handed children and those under 6 years or over 18 years.

Additionally, neurodevelopment image data from Healthy Brain Network (HBN) initiative were obtained as validation samples[76]. Given the transdiagnostic nature of the HBN database, stringent inclusion criteria were applied to maintain consistency with the discovery cohort. In addition to left-handedness and age restrictions, for the ADHD group, only participants with confirmed diagnoses meeting full criteria were included, while excluding those with IQ≤70, history of mood or psychotic disorders, or prior psychotropic medication use.

To perform retrospective image quality checks, the Computational Anatomy Toolbox (CAT12) was used to automatically identify problems in each T1-weighted image (e.g., noise and intensity inhomogeneities) and provide a weighted overall image quality rating (IQR)[77]. Only images with IQR<3.5 were retained in the final sample to meet CAT12’s threshold for satisfactory quality (rating scores=70).

### 4.2. Image Data Preprocessing

We generated individual GMV maps derived from anatomic T1-weighted images by using the CAT12 pipeline and the Statistical Parametric Mapping (SPM12) toolbox[77]. During voxel-based morphometry analysis, we segmented individual images into probability maps of gray matter, white matter, and cerebrospinal fluid tissue, then normalized the gray matter maps into the Montreal Neurological Institute standard space (MNI) through Diffeomorphic Anatomical Registration Exponentiated Lie Algebra (DARTEL) pipeline[78]. Voxel-wise GMV maps were then produced following bias-field correction and modulation using both linear and nonlinear components of the Jacobian determinant[15], and we employed spatial smoothing with 8mm full-width at half-maximum Gaussian kernel to improve the signal to noise ratio of GMV estimates. The Automated Anatomical Labeling atlas, an extensively used neuroanatomical atlas, was adopted to parcel the whole brain gray matter into 90 regions[27].

### 4.3. Construction of Morphometric Similarity Networks

To quantify the morphometric similarity patterns among brain gray matter regions (Figure 1b), we employed the KLS metric, a robust analytical approach for assessing the concordance of gray matter volume distributions between paired anatomical structures[79, 80]. The probabilistic relationships between regional pairs were derived through kernel density estimation, which generated probability density functions for subsequent KLS computation. To optimize computational efficiency while maintaining statistical reliability, we implemented a sampling framework comprising 2^7^ points. The KLS values range from 0 to 1, with higher values indicating stronger morphological similarity between regions and shorter edges in the covariance network[81]. This procedure resulted in a 90 × 90 matrix quantifying the structural similarity of gray matter for each subject.

### 4.4. Characterization of Topological Properties

Each individual’s GMV-derived covariance network comprises 90 parcellated regions, connected by edges that indicate their morphometric coupling[82]. We calculated three topological metrics—DC, NE, and PC to comprehensively investigate the hub organization of this network (Figure 1c). For each node, DC measures the number of edges to which it is connected, NE measures the average inverse shortest path length that links it to all other nodes, and PC quantifies how evenly its connections are distributed across different network modules (determined through greedy optimization algorithm, detailed definitions and calculations of these metrics can be found in the Supplementary Methods). Using the GRETNA toolbox[83], we applied a sparsity range of 0.10 to 0.34 with increments of 0.01 to generate sparse networks with minimal spurious edges, aligning with a small-world regime. To characterize the connectome independent of arbitrary sparsity thresholds[84, 85], we estimated the area under the curve for each of the three nodal topological phenotypes.

### 4.5. Normative Modeling

Normative models for each topological phenotype were implemented using the Bayesian linear regression framework from PCNtoolkit[10]. This approach estimates normative ranges for target phenotypes (i.e., the topological metrics) with respect to covariates of interest (i.e., age and sex) in the training dataset (Figure 1d). The degree to which individuals in a held-out test dataset deviate from these normative expectations is then quantified using *Z*-scores. Prior to model fitting, we excluded subjects with non-fitting outliers (|*Z*|>7) and brain phenotypes were subjected to Yeo-Johnson transformations to mitigate skewness[15]. Our datasets were then strategically partitioned: 90% of typically developing controls (TDC) subjects from each site were allocated to the training dataset, while the remaining 10% of TDCs, together with all ADHD cases, constituted the test dataset to examine whether ADHD group have more pronounced deviations than TDC. Lastly, Bayesian linear regression was employed to estimate region-specific normative variance between covariates and deviations for each topological metric, and to model uncertainty. These models incorporated demographic covariates (age and sex) and site-specific effects, utilizing B-spline basis expansion for age and likelihood warping to accommodate non-Gaussian nonlinear effects (detailed settings and basic principles of normative model provided in Supplementary Methods)[86]. Separate normative models were fitted for each of the three topological metrics in each of 90 brain regions. Site-related artifacts were assessed through a two-fold linear support vector machine classifier, yielding mean balanced accuracy across sites, groups, and metrics to validate data harmonization[15].

### 4.6. Characterization of Extreme Deviations

Individual ADHD deviation patterns were quantified by positioning their nodal topological metrics along normative centiles derived from TDC of the training data, and *Z*-scores were computed to characterize the magnitude and direction of deviations. For TDCs, corresponding deviation patterns were established using 10-fold cross-validation. To evaluate model robustness and generalizability for each brain region across topological metrics, we computed explained variance, mean squared log-loss, and standardized mean squared error[87]. To quantify nodal heterogeneity, we identified extreme deviations in topological patterns (|Z|≥2.0)[30, 31] and calculated the proportion of children showing atypical increases or decreases at each node. ADHD deviation maps were then subtracted from those of TDCs to obtain maps of case-control differences in extreme deviations. To assess the statistical significance of these differences, we performed both group-based and spatial permutation tests with 5,000 iterations to generate null distributions of case-control differences (details about random assignment of group labels and surrogate deviation maps provided in Supplementary Methods), with *p*-values calculated as the proportion of null differences that exceeded the empirical differences[15]. We identified nodes with statistically significant differences using thresholds of both *p*_uncorreced_<.05 and *p*_FDR_<.05 (two-tailed, Benjamini-Hochberg procedure).

### 4.7. Multimodal Fusion Framework

We implemented mCCA + jICA to topological phenotypes using the Fusion ICA Toolbox[32, 54], as a data-driven approach to reveal co-varying multimodal topological patterns with significant case-control differences (Figure 1e). Briefly, the analytical framework employed a two-stage process: initially, mCCA identified inter-modal relationships by optimizing correlations among loading parameters (X: topological vectors, Y: derived canonical variates), then jICA delineated spatial maps to elucidate case-control variations (details presented in Supplementary Methods)[88]. The minimum description length criterion was used to determine the optimal number of components[89]. The joint decomposition yielded modality-specific components and corresponding individual-specific mixing coefficients (*A_k_*, *k*=1,2,3). The independent sources (*S_k_*) for each modality were derived from the concatenated maps and corresponding whitening matrix, and we normalized independent sources into *Z*-scores and then thresholded at |*Z*|≥2.0 to visualize spatial maps per topological modality. To assess clinical relevance, we compared mixing coefficients (*A_k_*) between groups using two-sample *t* tests to identify joint group-discriminative component, and then evaluated their associations with clinical measures by product-moment correlation analysis.

Given that subscale scores measuring these phenotypic dimensions have demonstrated robust convergence and content validity[90], we standardized ADHD symptom assessment across sites to facilitate cross-site individual predictions. Symptom subscales were rescaled to a range of 0.0-1.0 (termed normalized ADHD symptom index), with each score calibrated relative to its site-specific maximum value[91, 92].

### 4.8. Biotype Identification

Unlike conventional unsupervised clustering algorithms (e.g., k-means), the semi-supervised HYDRA algorithm leverages a linear support vector machine to establish multiple hyperplanes that simultaneously encompass the control distribution while optimally differentiating it from the cases, thereby serving dual purposes of classification and clustering. Through the integration of these hyperplanes, the algorithm constructs a convex polytope, wherein each facet represents a distinct biotype (Figure 1f).

To capture the heterogeneity of brain topological properties, we trained HYDRA models utilizing a 10-fold nested cross-validation protocol to determine the optimal *k*-dimensional space. Specifically, the analysis was constrained to *k* = 2 to 4, as higher dimensionality was deemed impractical in clinical settings of ADHD. The clustering stability was quantified through the ARI. Details on HYDRA settings and reproducibility analyses are provided in Supplementary Methods. The robustness of the identified ADHD biotypes was validated via three reproducibility analyses. First, we conducted permutation testing to generate null distributions and compared their corresponding ARIs with those derived from a null model generated by randomly assigning TDC subjects into pseudo-groups to determine the statistical significance for the optimal *k*-cluster solution[93]. Second, since ADHD is more commonly diagnosed in boys, we also repeated our clustering analysis after excluding female participants, to account for any sex-specific effects. Finally, we executed a split-half cross-validation with 20 random iterations to assess if biotypes in each half showed similar deviations for the optimal *k* clustering, incorporating stratification by diagnosis, sex, and site[33].

To investigate the corresponding clinical profiles of the ADHD biotypes, Kruskal-Wallis analysis was used to examine their differences in symptom severity (i.e., inattention and hyperactivity/impulsivity phenotypes) in a dimensional approach[39]. Similarly, to quantify differences in atypical topological patterns among biotypes, we computed the frequency of extreme deviations within each node and used chi-square analyses to assess statistical differences in these distributions across the three biotypes (both *p*_uncorreced_<.05 and *p*_FDR_<.05 as two-tailed significance thresholds). Biotype-specific extreme deviation maps were depicted using the same procedures described above by computing the proportion of children with suprathreshold deviations, and we also performed group-based permutation tests to assess their statistical significance compared to patterns of TDCs.

To examine longitudinal changes in ADHD symptoms across biotypes, we analyzed follow-up data from the Child Behavior Checklist collected annually over four years at the WCH site (exclusively beginning with medication-naïve participants) using linear mixed-effect modeling[94]. The models incorporated time, biotype, and their interaction as fixed effects, with age and sex as covariates. To account for within-subject correlations in repeated measurements, we specified random intercepts for each participant. We fitted the model using the Newton-Raphson method (maximum iterations: 1,000; convergence tolerance: 1×10^−8^) and set statistical significance at *p*<.05. Additionally, at the WCH site, we monitored the development of mood disorders (e.g., anxiety and depression) among ADHD participants through KSADS interviews during follow-up. Chi-square analysis was used to assess differences in mood disorder comorbidity rates between biotypes.

### 4.9. Biological Correlates of Biotype Profiles

To elucidate molecular signatures underlying our topology-derived ADHD biotypes, we characterized their correspondence to the spatial distributions of neurotransmitter receptors (Figure 1g)[36]. The neurotransmitter receptor maps encompass density distributions of 19 distinct receptors, categorized into nine major neurotransmitter systems: serotonin, dopamine, acetylcholine, histamine, glutamate, norepinephrine, GABA, opioid, and cannabinoid, derived from PET imaging studies (details of included PET studies are provided in Supplementary Table S13)[37]. Group-averaged receptor maps from individual studies were registered to a standard MNI template and parcellated based on the AAL atlas. In cases where multiple tracers targeted the same receptor, we used a weighted averaging procedure and normalized each receptor map across brain regions to obtain relative density distributions[36]. To comprehensively characterize brain network organization of ADHD, we aggregated case-control difference maps in extreme deviations across three topological metrics (termed as fused topological deviations), capturing the network hubness based on diverse connectivity properties[28]. We then used Spearman correlation to quantify the spatial correspondence between integrating topological deviations (maps in unimodality as well) and neurotransmitter receptor distributions[95]. Statistical significance was assessed using spin tests, which accounted for spatial autocorrelation through comparison with 5,000 surrogate deviation maps[96].

We then used the Neurosynth-based meta-analytic task activation maps to contextualize our findings beyond the available clinical measures and to explore associations with a broader spectrum of psychological processes[38]. Neurosynth Compose (https://compose.neurosynth.org/), an automated neuroimaging meta-analytic platform, leveraged 30,578 fMRI studies (as of November 11, 2024, sourced from the NeuroStore repository) to derive meta-analytic task activation maps using coordinate-based multi-kernel density analysis (Chi-square workflow with association test maps) with FDR correction[97]. We selected activation maps corresponding to 123 cognitive terms, identified through their intersection with the Cognitive Atlas framework (as described in Hansen et al.)[98, 99], and these terms encompassed a comprehensive range of neurocognitive processes. The maps were subsequently parcellated and normalized to construct a region-by-cognitive function matrix for each term. Neurosynth-derived probabilistic activation maps provided quantitative representations of region-specific neural activity patterns associated with distinct psychological processes.

Partial least squares (PLS) regression analysis was employed to examine the spatial relationship between maps of fused topological deviations (dependent variable) and cognitive terms (predictor variable) across biotypes, with spin tests to correct for spatial autocorrelation (5,000 iterations)[95]. We then assessed the stability of cognitive term weights on the most significant PLS component with highest explained variance using a bootstrapping approach (10,000 iterations). The relative contribution of each cognitive term was quantified by calculating *Z* scores based on the ratio of term weights to their bootstrap-estimated errors. Terms were subsequently ranked according to their PLS component contributions, with significance determined by both spin-derive *p*-values and FDR-corrected thresholds (spin tests based on spherical rotations, *p*_spin_<.05 and *p*_spin-FDR_<.05). Lastly, Neurosynth decoding also serve as a valuable validation tool for our biotype categorization by examining whether the Neurosynth-derived cognitive terms (i.e., attention and hyperactivity/impulsivity) correspond to the clinical characteristics of each biotype.

### 4.10. Out-of-sample Validation of Biotypes

To assess the generalizability of our ADHD biotype clustering model, we transferred the pre-trained model to an independent validation dataset from the HBN. T1-weighted imaging data from HBN underwent identical processing steps as the discovery sample up to the normative model training stage. Since the HBN database was unseen in the initial training set, a transfer learning strategy was employed to fine-tune the reference model using TDC samples from HBN as adaptation data to accommodate potential site-specific variations[100]. During the adaptation process, hyperparameters of reference model were used as informed priors for the new dataset, and we adjusted the mean and variance within the latent Gaussian space before wrapping the adjusted data back to its original space[30]. Following recalibration of the reference normative range, we generated site-effect-free estimates of the validation sample to characterize their topological deviations. By leveraging HYDRA algorithm parameters (weight vector and bias term) that defined a set of hyperplanes during the initial training phase, we reconstructed the polytope that optimally distinguishes the TDC group from various ADHD biotypes and then applied it to ADHD validation data to compute expression scores across all dimensions (Figure 1h, detailed methodology available in the Supplementary Methods). ADHD subjects in the validation cohort were then assigned to clusters based on their maximum expression scores. Finally, we examined both clinical profiles and neural deviation patterns in the validation sample as we applied above to the discovery cohort.

## Acknowledgments

We express our gratitude to the participating families and clinical research coordinators for their contributions to the WCH and UC datasets. We acknowledge the ADHD-200, ABIDE, and HBN initiatives for their open-source pediatric neuroimaging data, which advances scientific research through open-science practices. We would also like to express our gratitude to Jianyu Li from the Huaxi MRI Research Center, West China Hospital of Sichuan University for his assistance with data extraction, and Ashlea Segal from the Department of Neuroscience, School of Medicine, Yale University for her assistance with model optimization.

## Author contributions

N.P., A.F., and Q.G. conceived and designed the study. R.K.M., M.P.D, Y.Chen, and Q.G collected a subset of the data for this study. N.P. and Y.L. performed the data analysis. N.P., Y.L., Q.C., and Z.Z preprocessed the data. Y.L., K.Q., Y.Cao, and A.F. contributed to interpretation of the results. N.P. and Y.L. wrote the manuscript. I.P., L.L., R.K.M., M.P.D, M.K.S, Y.Cao, and A.F. revised the manuscript. N.P. contributed to the visualization. All authors reviewed and approved the final manuscript.

## Data and Code Availability Statement

ADHD-200 and ABIDE datasets are available through repositories: https://fcon_1000.projects.nitrc.org/indi/adhd200/ and https://fcon_1000.projects.nitrc.org/indi/abide/, respectively. UC datasets could be accessed through the National Institute of Mental Health Data Archive (https://nda.nih.gov/) with approval from the principal investigator of University of Cincinnati. WCH datasets are only available upon reasonable request to the corresponding author due to the data policy and ethical restrictions. HBN initiative could be obtained from https://fcon_1000.projects.nitrc.org/indi/cmi_healthy_brain_network/.

The neuroimaging preprocessing toolbox is openly accessible (SPM12, https://www.fil.ion.ucl.ac.uk/spm/software/spm12/; CAT12 https://neuro-jena.github.io/cat/). Toolbox that constructs MSN could be obtained from Dr. Jinhui Wang of South China Normal University. GRETNA toolbox could be downloaded at https://www.nitrc.org/projects/gretna/. PCNtoolkit could be obtained at https://github.com/amarquand/PCNtoolkit/. Fusion ICA Toolbox could be downloaded at https://github.com/trendscenter/fit/. The script that constructs HYDRA models could be obtained from https://github.com/evarol/HYDRA/. Neurotransmitter distribution maps could be downloaded at https://github.com/netneurolab/hansen_receptors/. Neurosynth-based associated maps of cognitive term could be derived from Neurosynth Compose (https://compose.neurosynth.org/). Brain charts were visualized by MRIcroGL (https://www.nitrc.org/projects/mricrogl) and Surf Ice (https://www.nitrc.org/projects/surfice/). All other associated code and pre-trained models to reproduce the analysis results of normative models, fusion models, prediction of HYDRA clustering, and neural contextualization will be openly available at https://osf.io/wjgsr/ upon publication of the article. All other data supporting our findings are provided within the paper and supplementary information.

